# Whole-Blood DNA Methylation Analysis Reveals Respiratory Environmental Traits Involved in COVID-19 Severity Following SARS-CoV-2 Infection

**DOI:** 10.1101/2021.11.03.21260184

**Authors:** Guillermo Barturen, Elena Carnero-Montoro, Manuel Martínez-Bueno, Silvia Rojo-Rello, Beatriz Sobrino, Clara Alcántara-Domínguez, David Bernardo, Marta E. Alarcón-Riquelme

**Affiliations:** GENYO. Center for Genomics and Oncological Research Pfizer/University of Granada/Andalusian Regional Government. Granada, Spain; Servicio de Microbiología e Inmunología. Hospital Clínico Universitario de Valladolid. Valladolid, Spain; Servicio de Enfermedades Infecciosas. Hospital Regional de Málaga. Málaga, Spain; Lorgen G.P., S.L., Business Innovation Center - BIC/CEEL, Technological Area of Health Science, Granada, Spain; Mucosal Immunology Lab. Unidad de Excelencia Instituto de Biomedicina y Genética Molecular de Valladolid (IBGM, Universidad de Valladolid-CSIC). Valladolid, Spain; Centro de Investigaciones Biomédicas en Red de Enfermedades Hepáticas y Digestivas (CIBERehd). Madrid, Spain; Unit of Inflammatory Chronic Diseases, Institute of Environmental Medicine, Karolinska Institutet, Stockholm, Sweden

**Keywords:** EWAS, SARS-CoV-2, COVID-19, Epigenomics, mQTLs, Autoimmunity

## Abstract

SARS-CoV-2 causes a severe inflammatory syndrome (COVID-19) leading, in many cases, to bilateral pneumonia, severe dyspnea and in ∼5% of these, death. DNA methylation is known to play an important role in the regulation of the immune processes behind COVID-19 progression, however it has not been studied in depth, yet. In this study, we aim to evaluate the implication of DNA methylation in COVID-19 progression by means of a genome-wide DNA methylation analysis combined with DNA genotyping.

The results reveal the existence of epigenomic regulation of functional pathways associated with COVID-19 progression and mediated by genetic loci. We found an environmental trait-related signature that discriminates mild from severe cases, and regulates IL-6 expression via the transcription factor CEBP. The analyses suggest that an interaction between environmental contribution, genetics and epigenetics might be playing a role in triggering the cytokine storm described in the most severe cases.

## Introduction

SARS-CoV-2 virus infection has affected millions of people during the last year worldwide. Most infected SARS-CoV-2 individuals remain asymptomatic or with mild symptoms that do not require hospitalization (∼81%), while in other, the virus cause a severe inflammatory syndrome called COVID-19 that primarily affects the lungs leading, in many cases, to bilateral pneumonia, severe dyspnea and in ∼5% of the cases, death^1,2^.

Several genetics, transcriptomics, and proteomics molecular studies have been performed to date, disentangling important pathogenic molecular mechanisms of the disease (3-14). In summary, SARS-CoV-2 infects the cells expressing surface receptors ACE2 and TMPRSS2^3^ causing cell damage due to its replication and release from the host cell. Then, this process triggers in the surrounding cells the production of pro-inflammatory cytokines and chemokines (including IL-1, IL-6, IL-8, IL-10, TNF and interferon inducible molecules, among others), which establish a pro-inflammatory response mediated by the accumulation of specific immune cells^4^. In severe cases, an overexpression of cytokines is produced in lung tissues, known as cytokine storm, thus provoking an over-response of the immune system and causing tissue damage. In the most critical cases, the cytokine storm is spread to other organs leading to multi-organ failure and death. Currently, the molecular mechanisms and the pathophysiology behind COVID-19 progression are largely studied and well established, but it is still unclear what makes some individuals develop the severe illness. In this sense, underlying genetic variation^5^ and different comorbidities have been identified as risk factors, such as diabetes, hypertension, chronic lung disease or even neurological disorders^6,7^. Also life style habits, that might be causing the previous conditions have been also related to COVID-19 illness as obesity or smoking, as well as age, gender or ethnicity^8,9^. However, it is unclear how these comorbidities, environmental and demographic conditions together with genetics, predispose and regulate the molecular mechanisms behind COVID-19 severity.

In order to shed light into the molecular relationship between risk factors and the regulation of the mechanisms behind the COVID-19 severity, here we present a DNA methylation EWAS (epigenome wide association analysis) combined with DNA genotyping for 473 and 101 SARS-CoV-2 lab positive and negative tested individuals recruited in two independent clinical centers. In addition to the study of the epigenetic regulation of COVID-19 pathogenic mechanisms, the DNA methylation changes associated with COVID-19 progression, and its genetic regulation were put in context by comparing the results with DNA methylation changes occurring in systemic autoimmune diseases (SADs), and with GWAS (genome wide association analysis) and EWAS catalogs that collect multiple traits described as potential COVID-19 severity risk factors.

## Results

### COVID-19 severity relates to impaired blood cell proportions and epigenetic activation of the innate immune response

Main blood cell type proportions were deconvoluted from the methylomes, showing a significant increase of neutrophil proportions associated with severity of the disease (Figure 1A and Supplementary Figures 1A and 1B). This imbalanced neutrophil proportion has been already shown to be related to COVID-19 severity progression ^10^, and proposed as an early prognostic signature ^1^. Besides cell proportion differences, significant differences in age and gender between groups were found in the discovery dataset (Wilcoxon test *p*-value < 0.05 for age in severe group compared to mild and negative individuals, and Fisher’s exact test *p*-value < 0.05 for gender proportion in severe group compared with mild group). Methylation plates did not show batch bias, being the largest bias between cohorts (Supplementary figures 1B and 1D). Based on these results, differential methylation analyses included as covariates: gender, age and the six major deconvoluted cell proportions.

**Figure 1:**
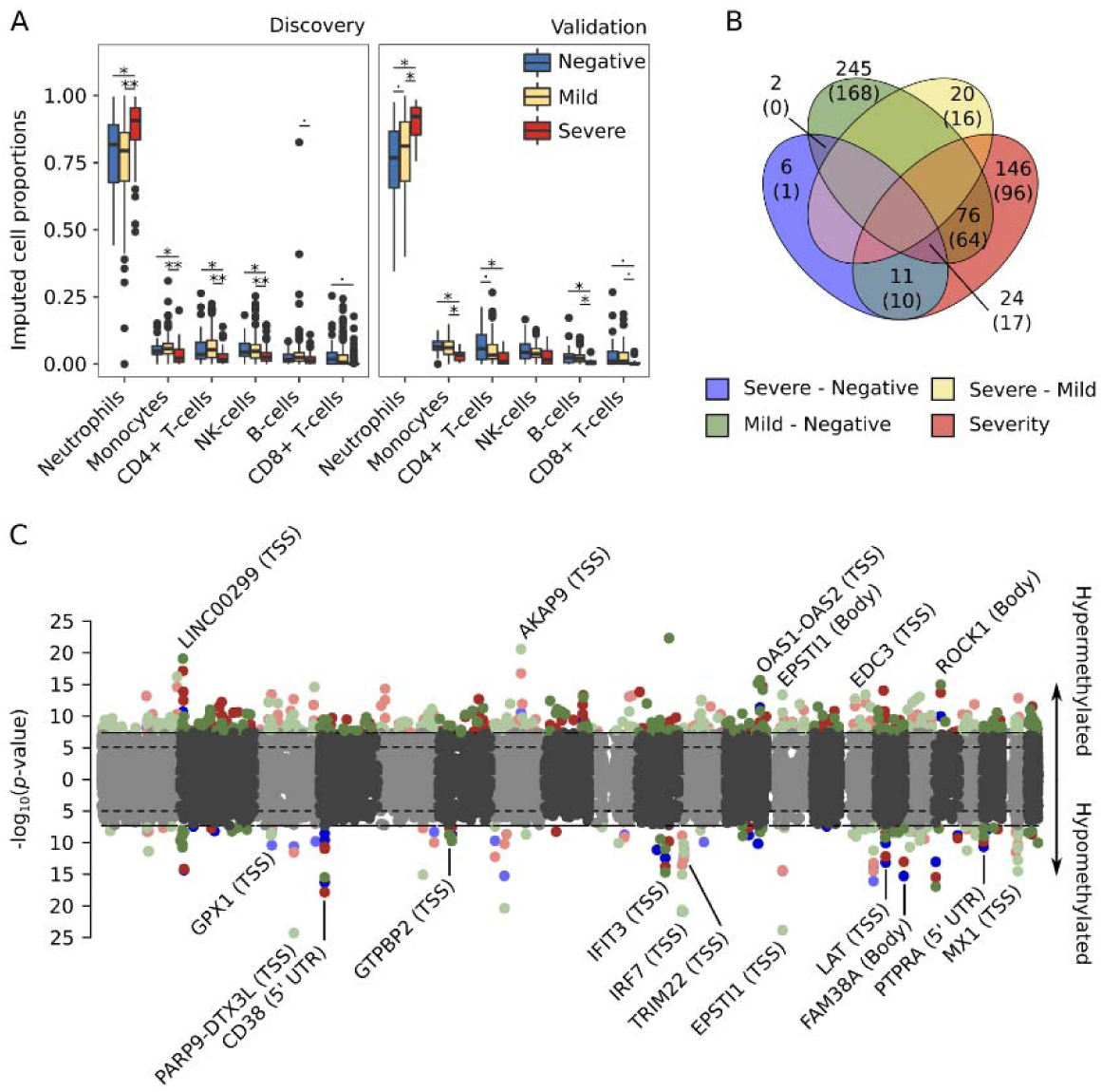
COVID19 severity correlates with an increase in blood neutrophil proportion and epigenetic changes in genes related with the innate immune response. (A) Methylome deconvoluted blood cell proportions are plotted by cohort (left panel discovery, right panel validation) and severity group (blue, negative SARS-CoV2 lab tested individuals; yellow, positive individuals with mild symptoms and red, positive individuals with severe symptoms). Paired differences were assessed by means of linear regression analysis (age and gender were included as covariates) and significance values plotted by pairs (*p*-value < 0.05, ^*^*p*-value < 0.01 and ^**^*p*-value < 1e-5). (B) Venn diagram with the number of significant shared DMCs across the differential analysis performed (the number of annotated genes are included in parentheses). (C) Combined manhattan plots are shown for the differential analysis that share DMCs, hypermethylated and hypomethylated DMCs are divided into upper and lower side of the manhattan plot respectively. Genes annotated for the shared DMCs are depicted, including, in parentheses their co-localization with the annotated gene (TSS, Transcription Start Site: Body, gene body). Severe vs negative (blue), mild vs negative (green), severe vs mild (yellow) and pseudotime longitudinal analysis (red).

Differential analyses were performed by pairs and longitudinally, after translating groups’ severity to a numerical scale (severity analysis, hereafter). We identified 530 CpGs differentially methylated in at least one regression model, and replicated in the validation cohort. Out of these, 43 DMCs were found in the severe-negative comparison, 347 in the mild-negative, 20 in severe-mild and 257 in the severity analysis (significant DMCs can be consulted in the Supplementary Files). We observed high degree of sharing between DMCs obtained in different comparisons (Figure 1B), except for the severe-mild DMCs which did not overlap with any of the other analyses results’. These specific DMCs from the severe-mild analysis were hypermethylated in the severe condition. Overall, 24 DMCs, annotated into 17 different genes were shared between severe-negative, mild-negative and with the severity analyses (Figure 1B and 1C), which give a general idea of the epigenetic contribution to the progression of COVID-19. Most of the shared signatures are related to the activation of the viral defense type I interferon inducible genes (OAS1-OAS2 hypermethylated and PARP9-DTX3L, IFIT3, IRF7, TRIM22, MX1 hypomethylated), the hyperactivation of B and T lymphocytes (CD38, EPSTI1, LAT hypomethylated), and others, as EDC3, known to interact with ACE2 ^11^.

DMCs localization enrichment analysis showed that hypermethylated changes related to SARS-CoV-2 infection are more prone to occur outside CGIs and in introns and in enhancers for the hypomethylated sites (Supplementary Figure 2A). These genomic regions are known to be hot-spots of DNA methylation changes ^12^. However, most of the DMCs found in these analyses colocalize around the TSS (Transcription Start Site) and/or in the 5’-UTR of the nearest gene (Supplementary Figure 2B), due to the EPIC array probe selection. This probe’s preferential location facilitates the interpretation of the results, as hypermethylation and hypomethylation in 5’-end regions of the genes is directly related to the inactivation and activation of gene expression, respectively ^13^.

### COVID19 disease DNA methylation changes in neutrophils, B-lymphocytes and CD8+ T-lymphocytes regulate autoimmune and viral defense related functional pathways

Functional enrichment analyses based on *Reactome pathway database* was performed taking into consideration the groups compared and the direction of the effects. An enrichment of hypomethylated signals at interferon-inducible genes, herein called IFN signature, and enrichment of hypermethylated signals at genes involved in FCGR phagocytosis and CD209 signaling (DC-SIGN) was observed when positive SARS-CoV-2 are compared to negative SARS-CoV-2 individuals (Figure 2A). The activation of IFN signature genes is related with an active viral infection and in particular with SARS-Cov-2 infection ^10^. However, at DNA methylation level the impaired interferon response between mild and severe cases found at the transcriptional level ^14^ cannot be observed (Supplementary Figure 3). This suggests that the exhaustion of the interferon signature might be controlled at a different regulatory level.

**Figure 2:**
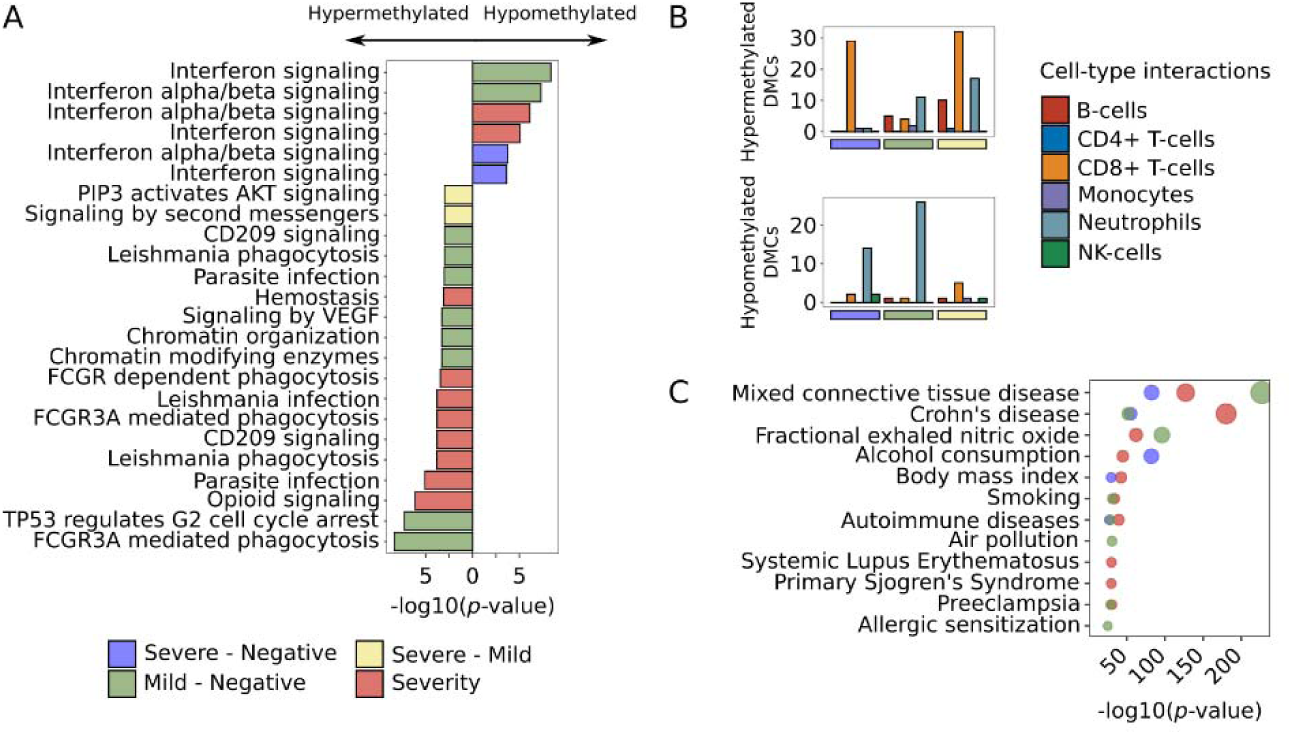
COVID19 DNA methylation changes regulate autoimmune related functional pathways and associate with environmental respiratory related traits. (A) Top 10 significant reactome database pathways (*p*-value < 0.01) are shown by differential analysis. (B) Number of DMCs with significant interactions for each deconvoluted cell-type proportion (red, B-cells; blue, CD4+ T-cells; orange, CD8+ T-cells; purple, monocytes; blue, neutrophils and green, NK-cells) are split into hypermethylated (upper panels) and hypomethylated (lower panel) and divided into the differential analysis. (C) EWAS traits enrichments (*p*-value < 1e-10) for each differential analysis are shown (MethBank database).

We performed interaction analysis between deconvoluted cell proportions and severity groups to identify which blood cell type is contributing to the epigenetic signatures. Our results suggest that interferon associated hypomethylation changes were mainly due to neutrophils and CD8+ T-lymphocytes (Figure 2B), while hypermethylation changes are primarily related to B-lymphocytes (Figure 2B) which in turn, might be related with the inactivation of CD209 signaling (Figure 2A). CD8+ T-lymphocytes also showed a number of significant hypermethylated interactions (Figure 2B) that may be related with the inactivation of FCGR3A phagocytosis-related genes in these cells (Figure 2A). Lastly, in the severe-mild analysis, methylation changes of the PIP3 activated AKT signaling pathway differentiate severe from mild COVID-19 patients (Figure 2A). Genes related with this pathway are hypermethylated in severe cases compared with mild COVID-19 cases, being CD8+ T-lymphocytes the major contributors to these changes (Figure 2B).

Finally, enrichment analyses were performed to assess to which other phenotypes or diseases the COVID-19 DMCs had been associated. For that, we used the information gathered in the *EWAS Atlas catalog* ^15^. Except for severe-mild DMCs, the other 3 comparisons showed DNA methylation changes in CpGs that were previously associated with different autoimmune conditions, allergy conditions, and an asthma related trait (as fractional exhaled nitric oxide test), but also with differential respiratory related environmental exposures (air pollution and polybrominated biphenyl exposure) and/or comorbidities that reflect lifestyle habits as body mass index, smoking or alcohol consumption (Figure 2C).

### Respiratory environmental related epigenetic changes differentiate severe and mild COVID-19 patients and mild COVID-19 cases from systemic autoimmune disorders

Significant DMCs from all the differential analyses performed were clustered together based on their methylation profile grouped by COVID-19 severity and divided into the two recruited cohorts (Figure 3A). Hierarchical clustering reveals that, aside from the significant values obtained in the linear regression models, not all trends of DMCs methylation changes are exactly replicated in both cohorts. Thus, 4 DMC modules were obtained based on the hierarchical clustering where DNA methylation changes were stable: S.Ho, composed by CpGs with a hypomethylation profile along COVID-19 severity; S.He, characterized by a hypermethylation profile along COVID-19 severity; M.Ho, in which hypomethylation events are observed in mild as compared with severe cases; and M.He, in which hypermethylation occurs in mild as compared with severe cases.

**Figure 3:**
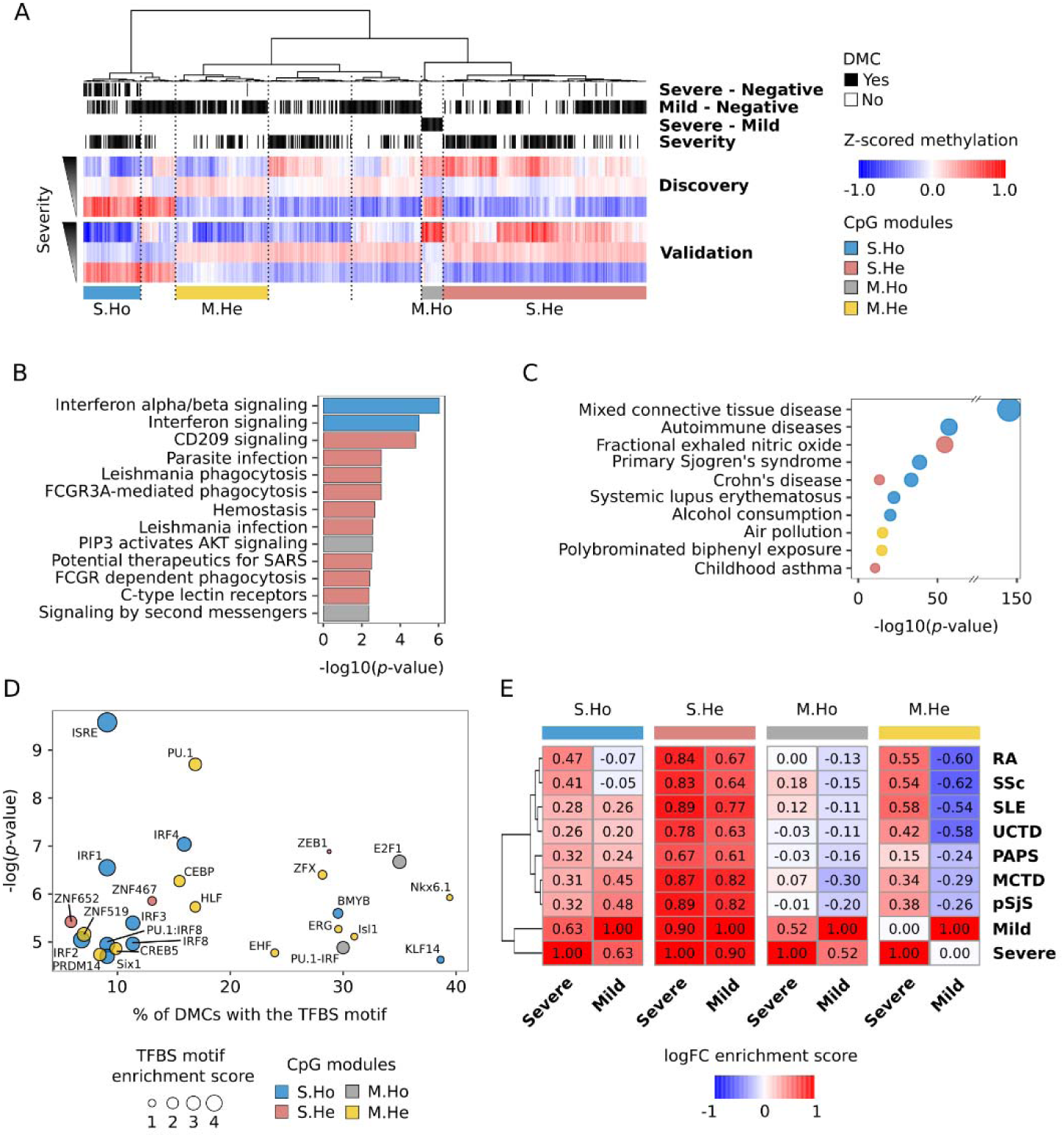
Epigenetic changes in CpGs associated with environmental respiratory traits differentiate COVID19 progression and mild cases from autoimmune disorders. (A) Hyerarchical clustering of methylation DMCs for both discovery and validation cohorts (Ward’s hierarchical agglomerative clustering with Pearson correlation as distance is used). Individual methylation values are averaged by severity from severe cases (top), mild cases (middle) to negative lab tested SARS-CoV2 (bottom). The annotations in the upper part of the plot refer to the analysis to which each CpG is differentially methylated (black). Four CpG modules highly replicated between cohorts, were selected from the hierarchical clustering: S.Ho (Hypomethylated with the severity), S.He (Hypermethylated with the severity), M.Ho (Hypomethylated in mild compared with severe patients) and M.He (Hypermethylated in mild compared with severe patients and healthy controls). (B) Reactome significant pathways by CpG module (*p*-value < 0.01) are shown. (C) MethBank EWAS trait enrichment by CpG module (*p*-value < 1e-10) are shown. (D) Significant overrepresentation of transcription factor binding site prediction (HOMER, *p*-value < 0.001) is depicted by CpG module. (E) Average log2FC Pearson correlations between COVID19 severity groups and seven different systemic autoimmune conditions (SLE, systemic lupus erythematosus; RA, rheumatoid arthritis; pSjS, primary Sjögren’s syndrome; SSc, systemic sclerosis; MCTD, mixed connective tissue disease; PAPs, primary antiphospholipid syndrome and UCTD, undifferentiated connective tissue disease). DMCs are grouped by CpG modules.

In summary, *Reactome* pathway enrichment analysis done on the 4 modules (Figure 3B) replicated the previous enrichments found for the DMCs grouped in the linear regression analysis (Figure 2A). Interestingly, a new additional pathway appeared to be enriched in the S.He module, related with potential therapeutics for SARS, which suggests that several of the proposed therapeutic targets for SARS infection are based on the activation of hypermethylated molecular pathways during the course of the COVID-19 disease. In order to validate the activation or inactivation of the enriched pathways revealed by means of the DNA methylation changes, *Reactome* pathways activity was estimated based on single-cell RNA-Seq information from publicly available analyses ^16,17^. The analysis was focused on the cell-types that mostly contribute to the DNA methylation changes: CD8+ T-lymphocytes, B-lymphocytes and neutrophils, as revealed from the interaction results (Figure 2B). In general, molecular pathway activities follow the DNA methylation changes at early sampling time points, which correspond to our recruited cohorts. This is, that pathways that show hypomethylation in certain group(s) of individuals coincide with a higher transcriptome activity compared with the hypermethylated groups, at least in the cell-types in which the change has been predicted to be occurring (Supplementary Figure 4). For example, the FCGR3A phagocytosis pathway (enriched in S.He module) activity is decreased with the severity of the disease in CD8+ T-lymphocytes, while the interferon signaling (enriched in S.Ho module) activity is increased with the severity. Certainly, at the transcriptome level, the interferon exhaustion signature associated with severe cases, not previously seen at the DNA methylation level (Supplementary Figure 3), can be appreciated for B-lymphocytes and CD8+ T-lymphocytes.

On the other hand, *EWAS Atlas catalog* enrichments were performed by modules, revealing that autoimmune and asthma related traits are mostly enriched in S.Ho and S.He modules, while the differential respiratory environmental related traits were mostly enriched in the M.He module. This M.He module differentiates severe and mild COVID-19 cases, suggesting an important contribution of the respiratory environmental exposure to the progression of COVID-19 disease, at least at the DNA methylation level.

TFBS motif analysis reveals specific TFBS motifs enriched for the different modules (Figure 3D). S.Ho module was mainly enriched in interferon regulatory TFBSs, in line with the *Reactome* pathway enrichment results, and among the other results stands out the enrichment of the CEBP motif in M.He module. CEBP is a transcription factor related with the inflammatory immune response by cooperating with and stimulating the transcription of different pro-inflammatory cytokines ^18^, among others, IL-6.

Given the potential relationship of COVID-19 affected molecular pathways and autoimmune disorders, DNA methylation profiles were compared between COVID-19 and the PRECISESADS collection ^19^, which includes DNA methylation information from seven SADs (Figure 3E). Both, severe and mild related DNA methylation changes correlate with systemic autoimmune disorders for S.He module, with a slightly higher intensity in severe COVID-19 patients. S.Ho module correlations are also significantly positive, except for RA and SSc comparison with mild cases, which present no significant correlations. RA and SSc patients are known not to be frequently expressing the IFN signature, enriched in S.Ho module ^20^. Thus, this result might be related with the presence of two signatures contributing to this module, one related with the interferon, which highly correlates with most interferon related SADs, and another one that correlates between severe, RA and SSc. In order to further investigate the differential correlation between SADs in this particular module, strongest hypomethylated CpGs in interferon related SADs and COVID patients (logFC < -0.25) corresponding with IFN signature genes, were removed from the correlation analyses (annotated in TRIM22-TRIM5, PARP9-DTXL3, RUNX1, IFIT3, IRF7, EPSTI1, MX1 and ADAR genes). The correlation without these CpGs shows a dramatic reduction for interferon related SADs, while RA and SSc correlations with severe cases are preserved (Supplementary Figure 5). This means that the remaining CpGs (annotated in genes as CCDC61, CD38, FAM38A, LAT, TREX1 or NFAT5, among others) differentially contribute to COVID19 progression similarities with SADs, some of them regulating the activation and differentiation of T and B lymphocytes. On the other hand, M.He module shows a strong correlation for severe and a strong anti-correlation with mild cases, thus differentiating mild cases from SADs. Lastly, M.Ho correlation results do not show significant correlation values.

### DNA methylation changes that differentiate mild and severe COVID19 cases show low genetic contribution and mQTLs enriched in SNPs associated with environmental traits

As the DNA methylation modules that mostly differentiate severe and mild cases (M.He) were mainly associated with environmental traits, we next interrogated whether there is genetic contribution behind these epigenetic changes, and how genetics contribute to the DNA methylation modules. In this sense, DNA methylation heritability was calculated for each CpG in the modules. Two independent methods showed high agreement in heritability calculation (Supplementary Figure 6A), so for the subsequent analysis variance decomposition model was selected. Genetic contribution to methylation variability was shown to contribute differentially between modules, being larger in S.Ho and S.He than in M.Ho and M.He modules (Figure 4A). This is in agreement with the higher environmental contribution to M.He shown by EWAS traits enrichments. Additionally, covariates as SARS-CoV-2 infection, age and gender were shown not to modify the genetic contribution to DNA methylation changes (Supplementary Figure 6B). S.Ho and S.He modules were the most affected by SARS-CoV-2 infection, while M.Ho and M.He variation might be driven by other covariates or environmental factors that, unfortunately, were not recorded in these cohorts (Supplementary Figure 6C).

**Figure 4:**
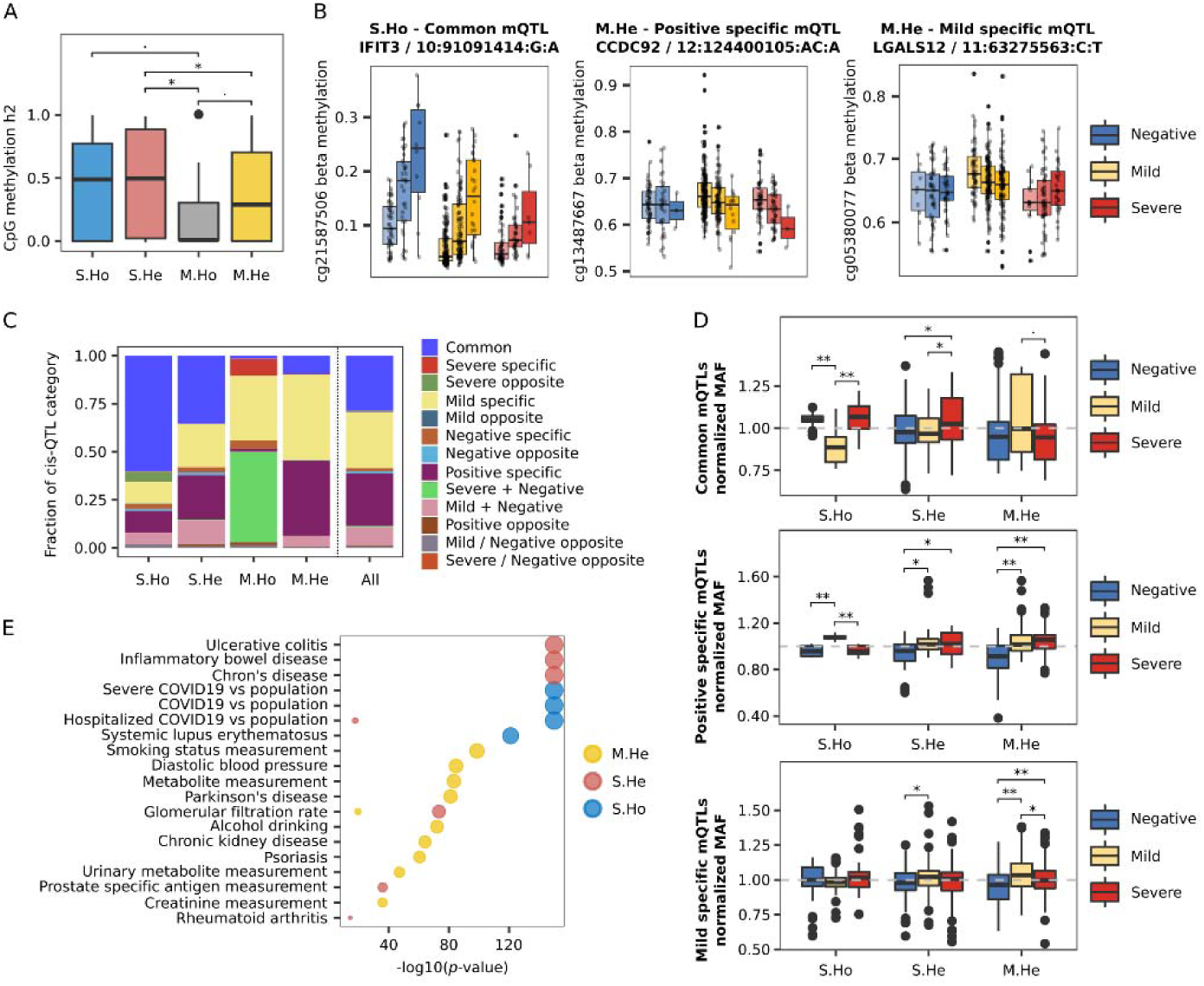
Genetics contributes differentially to progressive and mild specific DNA methylation changes. (A) Genetic contribution in terms of the fraction of the variance explained (heritability, h2) of individual CpG methylation changes is shown by DNA methylation module. Statistical differences are assessed by means of Wilcoxon test *p*-values. (B) Three significant mQTLs regulating DNA methylation levels are shown divided by severity group and genotype. From left to right, a common mQTL for all three severity groups in the S.Ho module, a positive specific mQTL and a mild specific mQTL for M.He module are depicted. (C) Fraction of mQTL categories are plotted by module and for all significant DMCs together. (D) Normalized MAFs for the largest mQTL categories (common mQTLs, positive specific mQTLs and mild specific mQTLs) represented in at least three modules (S.Ho, S.He and M.He) are shown divided by severity group. Wilcoxon test *p*-values were calculated between severity groups. (E) Enrichment of GWAS catalog and COVID-19 Host Genetics Initiative associated SNPs are shown by CpG module (*p*-value < 1e-10).

In order to further investigate the genetic contribution to the DNA methylation changes observed during COVID-19 progression, *cis*-mQTLs (methylation quantitative trait loci) were assessed (significant results can be consulted in Supplementary Files). Linear regression models were fit independently for each severity group (FDR < 0.05 for at least one group), showing that around 50% of the CpGs in each module were associated with at least one SNP (Supplementary Figure 6D). In total, 7899 unique mQTLs were found to be significant for at least one of the severity groups, composed of 7548 SNPs and 175 CpGs (out of 352 DMCs) with an average of 45±84 SNPs by CpG, what indicates that almost half of the DNA methylation changes found are being regulated by large blocks of SNPs in *cis*. mQTLs were classified according to the SNP-CpG association significance by severity groups, then labeling for example: a mQTL as mild specific, when the significant association (*p-*value < 0.05) was only found in COVID-19 mild cases, or positive specific, when both mild and severe cases showed a significant association (Figure 4B). mQTLs classification showed a differential genetic regulation by module (Figure 4C), where methylation changes, which follows COVID-19 progression (S.Ho and S.He modules), were enriched in mQTLs shared by all the severity groups (common mQTLs), which means that the genetic regulation of these DNA methylation changes does not depend on the severity of the disease but are a general regulatory mechanism. On the other hand, mQTLs in M.Ho and M.He modules were mostly identified as group-specific mQTLs, with a large fraction of mild and positive specific in M.He module, and mild, severe and severe/negative specific in M.Ho module. The genetic regulation specificity of M.He is also supported by significant differences of the normalized MAF (each group minor allele frequency divided by all groups’ minor allele frequency) for the mild and positive specific mQTLs (Figure 4D). MAF in positive specific mQTLs showed a higher frequency in mild and severe groups compared to negative individuals, while the mild specific mQTLs showed a higher MAF in mild cases. Surprisingly, MAF differences were found between mild compared to severe and negative individuals for common and positive specific mQTLs in S.Ho modules, which might indicate a differential genetic regulation also for mild individuals for the S.Ho signature (Figure 4D).

The enrichment of the significant mQTLs by module were tested for SNPs previously known to be associated with different traits. In this sense, mQTLs trait enrichments were performed considering the *GWAS catalog database* ^21^ and the COVID-19 associated SNPs from the COVID-19 Host Genetics Initiative ^5,22^. The results showed a strong enrichment of SNPs associated with COVID-19 and interferon related autoimmune diseases (Systemic Lupus Erythematosus) in the mQTLs regulating the S.Ho module and SNPs associated with non-interferon related autoimmune diseases in the S.He module (Figure 4E). On the other hand, M.He mQTLs were enriched with environmental related SNPs (Figure 4E), mimicking the enrichments shown above for the EWAS catalog. Interestingly, two different COVID-19 GWAS regions are regulating the S.Ho and S.He modules. In the case of the S.Ho module, its *cis*-mQTLs are composed of SNPs at 3p21.31 GWAS peak ^22,23^, found to be associated in severe, hospitalized and in general SARS-CoV-2 lab positive tested patients compared with the general population. While S.He module is enriched in SNPs located at 8q24.13 GWAS peak ^22^, only found to be statistically significant in hospitalized COVID-19 patients compared to general population ^22^.

## Discussion

The EWAS of SARS-CoV-2 infection reveals a DNA methylation regulation of important functional pathways related with COVID-19 progression and specific epigenetic differences between severe and mild patients. Differentially methylated CpG sites were shared between severe and mild cases, mainly associated with the activation of interferon signaling pathway and the hyper-activation of B and T lymphocytes. These pathways have been previously associated with COVID-19 severity in transcriptome studies ^10,24^, showing in this study that the regulation of these pathways is being mediated by epigenetic changes at the promoter level of the implicated genes (Figure 1).

Apart from the DMCs shared between the differential analyses, the pathways enrichment analysis for the individual regression models showed the epigenetic deregulation of specific pathways as CD209 signaling (DC-SIGN), FCGR phagocytosis pathway and AKT signaling in specific blood cell-types (Figure 2). CD209 is primarily expressed in dendritic cells and B-lymphocytes, and its interaction with CD209L, expressed in SARS-CoV-2 target tissue endothelial cells, has been shown to facilitate the virus entry ^25^. Thus, CD209 signaling hypermethylation might be playing a protective role during SARS-CoV-2 infection. Additionally, CD209 activation has been shown to promote B-lymphocyte survival ^26^. However, this process does not seem to be occurring in SARS-CoV-2 infection as shown by the B-lymphocyte depletion observed in the deconvolution analysis (Figure 1A). FCGR phagocytosis pathway is involved in the antibody-antigen complex clearance and the antibody dependent cellular mediated cytotoxicity. CD8+ T-lymphocytes expressing FCGR3A (CD16) have been described to acquire natural killer (NK) cell-like functional properties, thus contributing to their cytotoxic functionality, increased in chronic hepatitis C virus infections ^27^. Recently, suppression of cytotoxic activity has been described on CD8+ T-lymphocytes and NK-cells from severe COVID-19 patients ^28^, which in the light of our DNA methylation results might be impaired because of the DNA hypermethylation of genes of the FCGR3A phagocytosis pathway. Based on our results, these two pathways seem to be associated with the progression of the disease, showing significant DNA methylation changes along its course. On the other hand, gene promoters related with the AKT signaling pathway were specifically found to be differentially methylated when compared severe and mild cases (hypomethylated in mild), thus differentiating at the epigenome level severe from mild SARS-CoV-2 infected patients. AKT signaling in CD8+ T-lymphocytes is critical for the effector-memory transition of this cell-type ^29^, thus impairing the protective immune secondary response and potentially contributing to the worst outcome. Other important genes, not annotated in these pathways, were found to show methylation differences, as for example EDC3. Interestingly, hypermethylation of EDC3 in severe cases might be mediating the overexpression of ACE2 protein in SARS-CoV-2 patients, thus favoring infection ^3^. EDC3 is a component of a decapping complex that promotes removal of the monomethylguanosine (m7G) cap from mRNAs, being an important protein during mRNA degradation, and its interaction with ACE2 has been experimentally validated and shown with STRING interaction network ^30^.

In addition to the COVID-19 EWAS results, DMCs were grouped by hierarchical clustering and filtered by cohorts’ similarity (Figure 3). Four modules of co-regulated CpGs were found, where three of them are enriched in the functional pathways previously described. CD209 and FCGR phagocytosis pathways (S.He module) are hypermethylated with the severity of the disease, and both severe and mild cases, perfectly correlate with DNA methylation changes observed in SADs. Hypomethylation along COVID-19 severity module (S.Ho) was found to be composed by two signatures, an interferon related signature which correlates with interferon related systemic autoimmune diseases (as MCTD, SLE or pSjS) at both severe and mild cases, and a T and B lymphocyte activation signature, which correlates mainly with non-interferon related SADs (RA and SSc) for severe cases. The AKT signaling pathway was also represented in the mild hypomethylated module (M.Ho). The fourth module, hypermethylated in mild cases (M.He), is of particular interest. It perfectly discriminates between severe and mild COVID-19 cases, and severe DNA methylation changes are highly correlated with autoimmune conditions. Additionally, and in contrast to the other CpG modules, its CpGs have not been related with autoimmune conditions but with respiratory environmental conditions. Further analyses on this module revealed an enrichment in CEBP binding sites (Supplementary Figure 6C). CEBP transcription factor has an important role regulating IL-6 and IL-1 expression, whose elevated levels have been associated with severe complications of COVID-19 disease ^4^. This result shows a reduced activity of CEBP binding sites in mild cases compared with the severe ones, in a module where DMCs are enriched in respiratory environmental traits. Altogether, our results suggest the existence of a relationship between environmental exposure and the cytokine storm associated with the most critical outcomes of COVID-19 disease.

The genetic regulation of COVID19 associated DNA methylation changes were also studied, finding important differences between modules (Figure 4). In addition to a lesser genetic contribution to the DNA methylation changes in M.Ho and M.He modules, the mQTLs associated to these modules showed more group specificity than S.Ho and S.He modules. Importantly, *GWAS catalog* enrichments for the mQTLs showed again a predominance of environmental traits related SNPs for the M.He module, which reinforces the idea of the importance of the environmental exposure during the regulation of the DNA methylation changes in this module.

This study is the first in depth large EWAS comparing SARS-CoV-2 RT-PCR positive and negative individuals. The results show a large epigenetic regulation of autoimmune related functional pathways during COVID-19 progression that differentiate severe from mild COVID-19 cases. Some of these autoimmune related pathways presented DNA methylation differences between severe and mild cases with less genetic contribution, but with higher genetic specificity than changes that progress with the severity of the disease. Interestingly, these specific epigenetic changes were mainly related, in terms of DNA methylation sites and SNPs regulating these sites, with environmental traits. Thus, in the light of the results, the interaction between specific genetic changes and different environmental exposure or life habits might be deregulating, via DNA methylation changes, autoimmune related functional pathways which are related with the worsening of SARS-CoV-2 infection. Despite the relationship between environmental exposure and COVID-19 severity has been suggested in previous epidemiological studies, this is the first time that this relationship is supported by genetic and epigenetic molecular information, thus, contributing to the understanding of the disease at the molecular level. Of special importance is the association of these environmental related DNA methylation changes with the cytokine storm typical of the most severe COVID-19 cases.

## Methods

### Study design and cohorts

Whole blood samples from SARS-CoV-2 RT-PCR negative (101) and positive lab tested individuals (473) were obtained from two clinical centers (*Hospital Clínico Universitario de Valladolid, discovery cohort and Hospital Regional Universitario de Málaga*, validation cohort). The regional ethical committees from Andalucía (*Comité Coordinador de Ética de la Investigación Biomédica de Andalucía*) and from Valladolid (*COMITÉ DE ÉTICA DE LA INVESTIGACIÓN CON MEDICAMENTOS ÁREA DE SALUD VALLADOLID*) approved the protocols and gave their ethical approval for this study and all recruited individuals signed the informed consent prior to recruitment. Whole blood was sampled upon arrival to the emergency ward, within a week after first symptoms. Discovery and validation cohorts were recruited between March-April 2020 and August-October 2020, respectively. Positive individuals were divided into: severe (WHO 5-7), if they needed invasive respiratory support, ICU admission and/or died due to SARS-CoV-2 complications, and mild (WHO 2-4), if patients did not develop severe COVID-19 related symptoms. Severity groups between cohorts were gender balanced, but slightly significant differences were found in terms of age (Table 1).

**Table 1:** Cohorts’ demographic and clinical information. Number of individuals (#), age average ± standard deviation (Age), number and percentage of males (Gender), hospitalized individuals (Hospitalized) and deceased individuals (Deceased) are shown by severity group and cohort. (*) Discovery and validation cohorts showed significant differences in terms of age in mild and severe groups (Mann-Whitney U test *p*-value < 0.05) and also in terms of numbers of hospitalized mild symptoms patients (Fisher exact test < *p*-value 0.05).

| Discovery<br>(10 Technical Batches) |  |  |  |  |  | Validation<br>(3 Technical Batches) |  |  |  |  |
| --- | --- | --- | --- | --- | --- | --- | --- | --- | --- | --- |
|  | # | Age | Gender | Hospitalized | Deceased | # | Age | Gender | Hospitalized | Deceased |
| **Negative** | 47 | 63 ± 21 | 20 (43%) | - | - | 54 | 67 ± 20 | 27 (50%) | - | - |
| **Mild** | 269 | 67 ± 15* | 126 (47%) | 216 (80%)* | - | 91 | 61 ± 18* | 48 (53%) | 87 (96%)* | - |
| **Severe** | 98 | 76 ± 14* | 126 (47%) | 98 (100%) | 84 (86%) | 15 | 64 ± 18* | 126 (47%) | 15 (100%) | 10 (67%) |

### Genomic Analysis

#### DNA extraction

DNA was extracted from whole blood samples by means of the *QIAamp DNA Blood Mini* kit and the automatic platform *QIAcube Connect*. Afterwards, DNA quality was validated and normalized using the *NanoDrop 2000c* and the *Qubit4*.

#### Genotyping

DNA was normalized to 200-400ng and genotyped *with Illumina’s Infinium GSA-24*.*v3*.*0 BeadChip*, following manufacturer’s recommendations. Markers with genotyping rate > 99%, minor allele frequency > 1% and a *p*-value for Hardy-Weinberg Equilibrium > 1e-6 were selected. Samples showing genotyping rate < 98%, inconsistencies between reported and genetic sex and extreme heterozygosity values (−0.2 < Fhet < 0.2) were eliminated. The kinship coefficient was calculated for each pair of samples and one member of each pair with a value >= 0.2 was removed. Based on a set of Ancestry Informative Markers (markers which maximize the allelic frequencies across 1000Genomes populations), individuals with non-European ancestry components were eliminated. The resulting dataset from this quality control process was imputed in the Michigan Imputation Server ^31^, using Minimac4 and 1000Genomes as reference panel ^32^. After subsequent filtering of the imputation result we obtained a working dataset consisting of 504 samples and more than 9.5 million markers. Quality control of the genotyped data was performed with *Plink2*.*0*^33^.

#### Methylome profiling

DNA methylation information was profiled with the *Illumina’s Infinium MethylationEPIC BeadChip*, after sample normalization to 500ng and bisulfite conversion with *EZ-96 DNA Methylation Kit*, as recommended by the manufacturer. Methylomes were quality controlled by genotype concordance (>= 0.8) using shared SNP probes between platforms (genotypes were extracted after imputation but without post filtering), gender prediction agreement (outliers > 5 standard deviations), signal from noise detection *p*-value < 0.1 and minimum number of beads (>3) that passed the detection *p*-value, the last two criteria were applied for both probes and samples. Additionally, sexual chromosomes, cross-reactive probes and probes with overlapping SNPs from *dbSNP v*.*147* ^34^ were discarded. Methylation beta values were normalized by means of functional normalization. After quality control, 574 samples and 768,067 probes were selected. The entire process was performed with *minfi and meffil R packages* ^35,36^.

### Statistical Analysis

#### Deconvolution of cell proportions

Iterative hierarchical procedure implemented in *EpiDISH R package* ^37^ was used to estimate the main blood cell type proportions from methylome information with the robust partial correlation method ^38^. Whole blood cell type reference panel includes: neutrophils, monocytes, B-lymphocytes, CD4+ T-Lymphocytes, CD8+ T-Lymphocytes and natural killer cells.

#### Differential and interaction analysis

Differential methylation analyses were performed by linear regression models, including gender, sex and deconvoluted cell-proportions as covariates. Linear regression models including interaction terms between the groups of interest and deconvoluted cell proportions, were used to estimate the specific cell type(s) where the methylation changes occur, as proposed by Zheng et al ^39^. Methylation changes and interactions were considered significant at nominal *p*-values below 0.01 in discovery and validation datasets, and below a genome wide significant level of 5e-8 in the meta-analysis of both cohorts. Meta-analyses were performed with the restricted maximum likelihood (REML) method and fixed effects implemented in *metafor R package* ^40^.

#### Enrichment, correlation and co-localization analysis

DMCs (Differentially methylated CpGs) and/or genes that co-localized with them, based on the Illumina annotation (*ilm10b4*.*hg19 R package*), were analyzed. Functional pathway analysis were performed against *Reactome Pathway Database* ^*41*^ *using ReactomePA R package* ^*42*^. EWAS trait enrichments were tested within the *EWAS Atlas* database ^15^. PRECISESADS methylomes ^19^ from seven SADs (SLE, systemic lupus erythematosus; RA, rheumatoid arthritis; pSjS, primary Sjögren’s syndrome; SSc, systemic sclerosis; MCTD, mixed connective tissue disease; PAPS, primary anti-phospholipids syndrome and UCTD, undifferentiated connective tissue disease) and healthy controls were used to compare with COVID-19 epigenetic changes. TFBS (transcription factor binding site) motif enrichment analysis was performed with *HOMER software* ^*43*^ using a size of 200 nucleotides and including as background the CpGs interrogated with the EPIC array.

#### Molecular pathway activity analysis

Single-cell RNA-Seq datasets were obtained from *Schulte-Schrepping et al*. ^17^ (BD Rhapsody system dataset, including neutrophils) and *Ren et al*. ^16^ (10x Genomics chromium dataset, not including neutrophils). Cells from both datasets were selected based on: mitochondrial read percentage < 5%, hemoglobin read percentage < 1%, number of reads > 500 and < 6000, and number of genes profiled between 200 and 2000. After the quality criteria filtering, almost all non-neutrophil cells were lost from *Schulte-Schrepping et al*. dataset. Thus, CD8+ T-lymphocytes and B-lymphocytes were analyzed from the Ren *et al*. dataset and neutrophils from the *Schulte-Schrepping et al*.. Individuals were classified as early or late based on *Schulte-Schrepping et al*. definition (late, sampling >11 days after first symptoms) and authors ’defined cell-type annotation was used to select two subsamples of 2500 cells for each cell-type (500 cells per severity group and onset category). Molecular pathway activity values were estimated by means of *ssgsea* algorithm implemented in *escape R package* ^44^. HLA and Immunoglobulin genes were removed from the *Reactome* pathways before activity calculation.

#### Genetic statistical analyses

Overall genetic contribution to DNA methylation changes (heritability, h2) was estimated by means of two models: one based on variance decomposition analysis from a linear mixed-model ^45^ and the other one using the diagonalization trick ^46^. The kinship matrix for the former model was calculated by means of *popkin R package* ^47^, while for the diagonalization trick estimation, *gaston R package* recommendations were followed ^46^. Methylation quantitative trait loci (mQTLs) analyses were performed using the *matrix-eQTL R package* ^48^. We applied a linear regression model that tests the additive effects of allele dosages for each genetic variant on the DNA methylation levels, while correcting for age, sex, the deconvoluted cell proportions and the first two genetic principal components. We restricted analysis to *cis*-mQTL mapping (maximum distance between CpG and SNPs of 1Mb) and SNPs with minor allele frequencies (MAF) > 0.05. *cis-*mQTL analyses were performed independently on the different severity groups, using a FDR < 0.05 as significance threshold. Significant mQTLs were classified as common or specific QTLs based on whether the association nominal *p*-values were below 0.05 for all the severity groups or not. Then classifying non-common QTLs based on the severity groups that pass the threshold (QTL effects were took into consideration what might result on shared significant QTLs between groups but with opposite effects). mQTLs enrichments were tested against SNP associated traits from the GWAS catalog database ^21^ expanded with COVID-19 Host Genetics Initiative results ^5,22^. GWAS catalog traits were selected based on studies with a replication cohort and at least 50 SNPs below the genomic significant threshold (*p*-value < 5e-8). Traits annotation into mQTLs were performed based on linkage-disequilibrium blocks by means of *PLINK1*.*9 software* ^33,49^, applying *blocks* function ^50^ default parameters in a maximum window size of 1MB.

## Data Availability

Genotypes summary statistics can be accessed through COVID-19 Host Genetic Initiative web page (https://www.covid19hg.org/), included in project Determining the Molecular Pathways and Genetic Predisposition of the Acute Inflammatory Process Caused by SARS-CoV-2 (SPGRX). Methylation profiling will be available from Gene Expression Omnibus (GEO) upon manuscript publication.

## Data availability

Genotypes summary statistics can be accessed through COVID-19 Host Genetic Initiative web page (https://www.covid19hg.org/), included in project “*Determining the Molecular Pathways and Genetic Predisposition of the Acute Inflammatory Process Caused by SARS-CoV-2 (SPGRX)”*. Methylation data are available from Gene Expression Omnibus (GEO) at XXXXXXXX.

## Acknowledgements

This work has been supported through *Consejería de Transformación Económica, Industria, Conocimiento y Universidades* of the regional government of Andalucía cofounded by the European Union through European Regional Development Fund (FEDER, CV20-10150), *Consejo Superior de Investigaciones científicas* (CSIC-COV19-016/202020E155) and Junta de Castilla y León (*Proyectos COVID* 07.04.467B04.74011.0 and IBGM excellence programme CLU-2029-02). G.B. is supported by the Instituto de Salud Carlos III (ISCIII, Spanish Health Ministry) through the Sara Borrell subprogram (CD18/00153). The authors would like to particularly express their gratitude to the patients, nurses and many others who helped directly or indirectly in the consecution of this study.

## Author information

### Contributions

**MEAR and GB** designed the study; **SRR, BS and DB** recruited the patients, performed their clinical assessment, and obtained the samples; **CAD** performed genotyping and DNA methylation profiling; **MMB** performed genotype data curation, imputation and processing; **GB** performed DNA methylation data curation and processing; **GB and EC** performed data analyses; **GB, EC, MMB and MEAR** discussed and interpreted the results; and **GB and MEAR** wrote the entire manuscript. All authors approved of the content of the manuscript.

### Corresponding authors

Correspondence to Guillermo Barturen or Marta E. Alarcón Riquelme

## Supplementary Figures

**Supplementary Figure 1:**
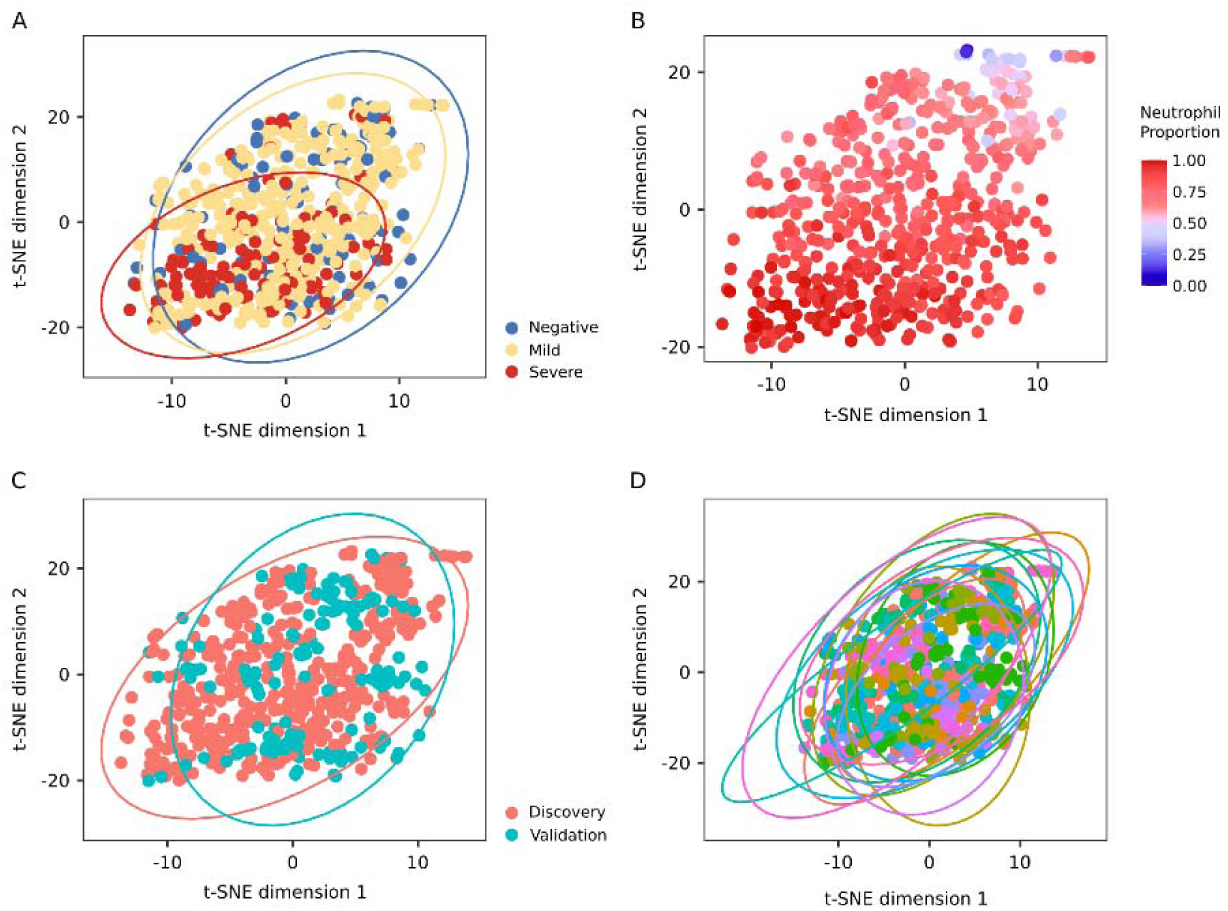
Technical batch effect does not bias the methylation profiles. t-SNE analysis of the 10.000 most variable CpGs (based on the DNA methylation absolute deviation mean) is shown colored by different variables: (A) severity groups, (B) neutrophil proportion, (C) cohorts and (D) technical batch.

**Supplementary Figure 2:**
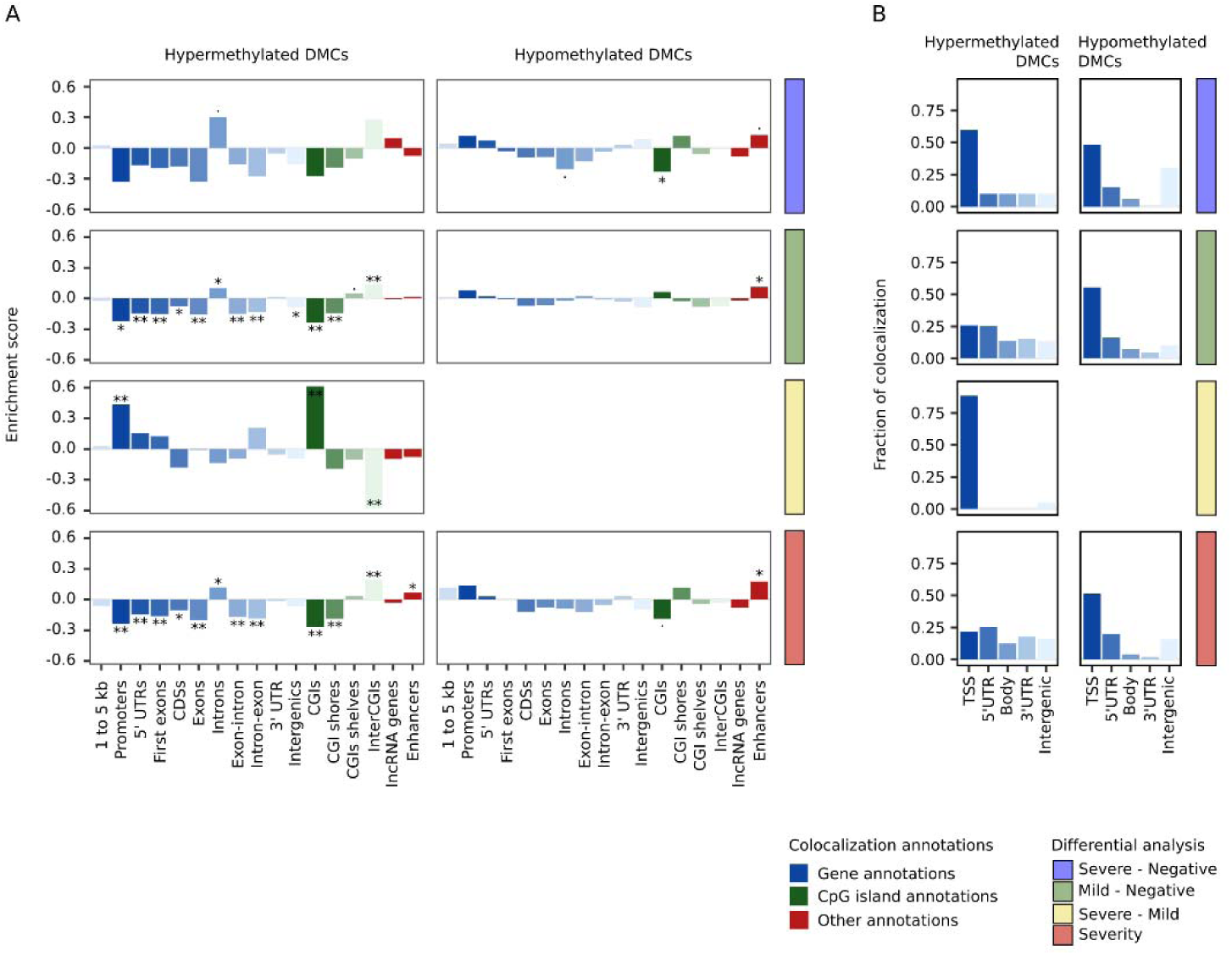
Hypomethylated and hypermethylated DMCs are mostly enriched and colocalized with gene regulatory elements, which tend to activate and inactivate in *cis* gene expression levels. (A) Significant DMCs enrichment from each differential analysis across different regulatory elements (annotatr R package). Hypermethylated and hypomethylated DMCs are divided into left and right panels, respectively. Each DMC is allowed to be annotated in more than one of the following features: 1 to 5kb, region between 1-5kb upstream from the TSS; promoters, region at less than 1kb upstream, from the TSS; 5’ UTR region; first exon; CDS, protein coding regions; exon; intron; exon-intron boundaries; intron-exon boundaries; 3’ UTR region; intergenic, not colocalized with any gene annotation; CGI, CpG island; CGI shores, at less than 2kb of a CGI; CGI shelves, at 2-4kb of a CGI; interCGI, not colocalized with any CGI annotion; lncRNA genes, GENCODE long non-coding gene annotation and enhancer, colocalized with FANTOM5 enhancer database annotation. Enrichment score is defined as the log2FC between the fraction of colocalized DMCs and the CpGs in the EPIC array. Significance was calculated by means of a Fisher exact test (*p*-value < 0.05, ^*^ *p*-value < 0.01 and ^**^ *p*-value < 1e-5). (B) Fraction of colocalized DMCs by differential analysis for ranked gene features obtained from the EPIC array annotation (each DMC is assigned to one feature according to: TSS, transcriptions start site > 5’ UTR > 3’ UTR > Body, gene body not in the previous features > Intergenic, not assigned to any gene).

**Supplementary Figure 3:**
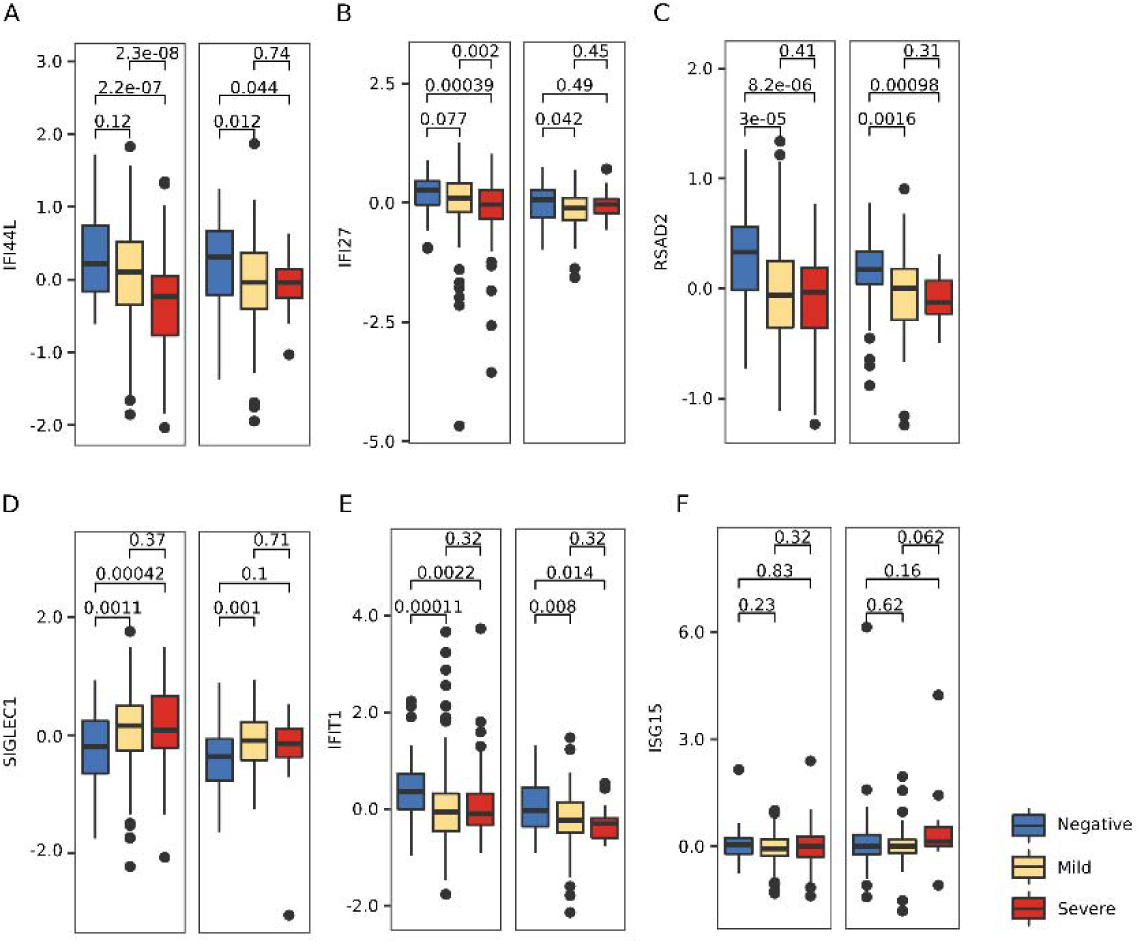
Interferon exhaustion in severe COVID-19 patients is not regulated by DNA methylation changes. (A-F) DNA methylation z-scored levels for CpGs colocalized with interferon gene signature promoters are shown by COVID severity group in discovery and validation cohorts. Wilcoxon test *p*-values are depicted by pairs.

**Supplementary Figure 4:**
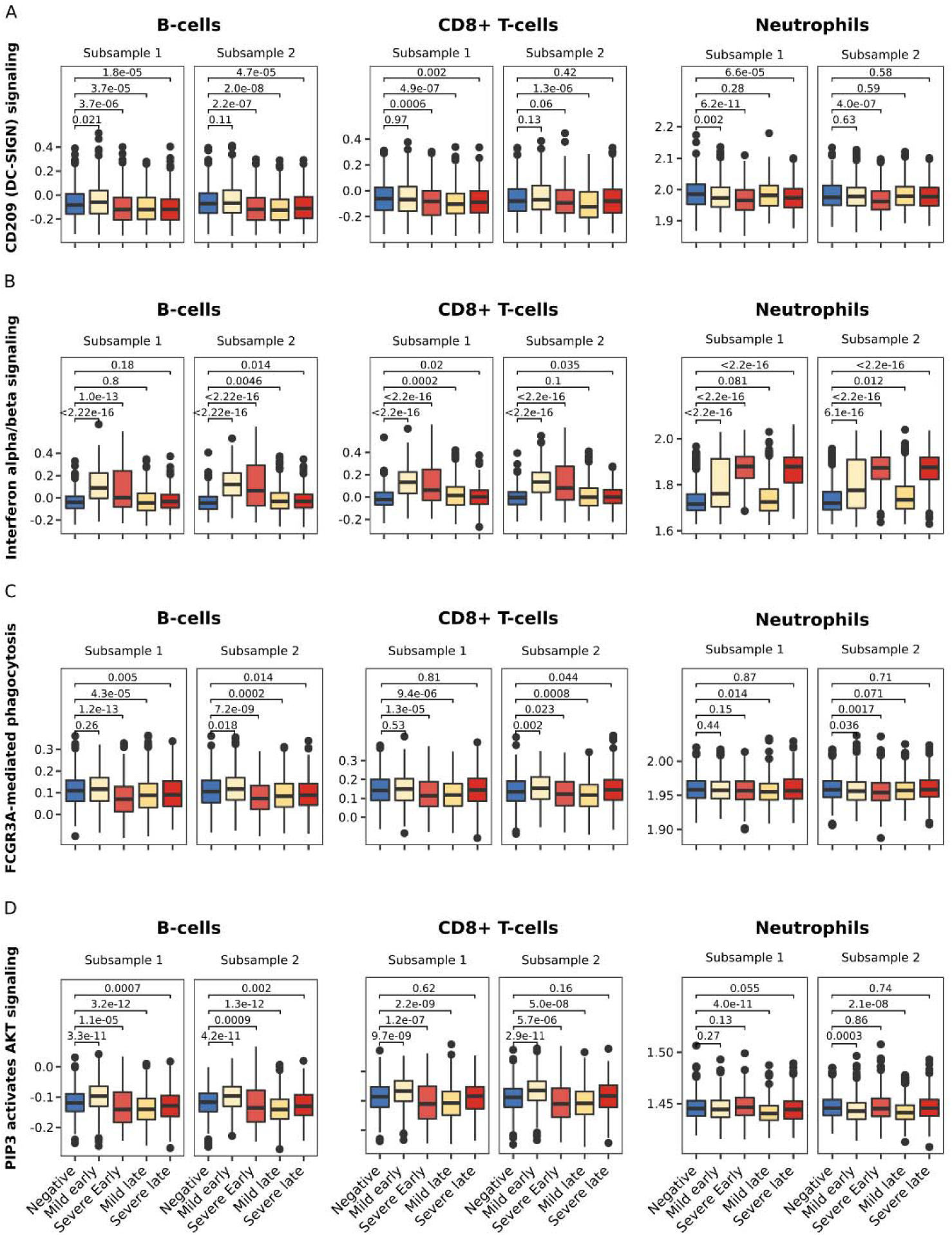
Enriched pathway activity in the CpG modules follow DNA methylation changes at early SARS-CoV-2 samplings in the cell-types with significant interactions. Reactome CD209 signaling (A), interferon alpha/beta signaling (B), FCGR3A-mediated phagocytosis (C) and PIP3 activates AKT signaling (D) activities were calculated per individual with ssgsea R package and grouped by COVID-19 severity groups at early and late samplings (>11 days after first symptoms) for B-cells, CD8+ T-cells and Neutrophils. Activities were plotted for two randomly selected subsets of 2500 cells, 500 cells per group. Wilcoxon test *p*-values are depicted against healthy controls.

**Supplementary Figure 5:**
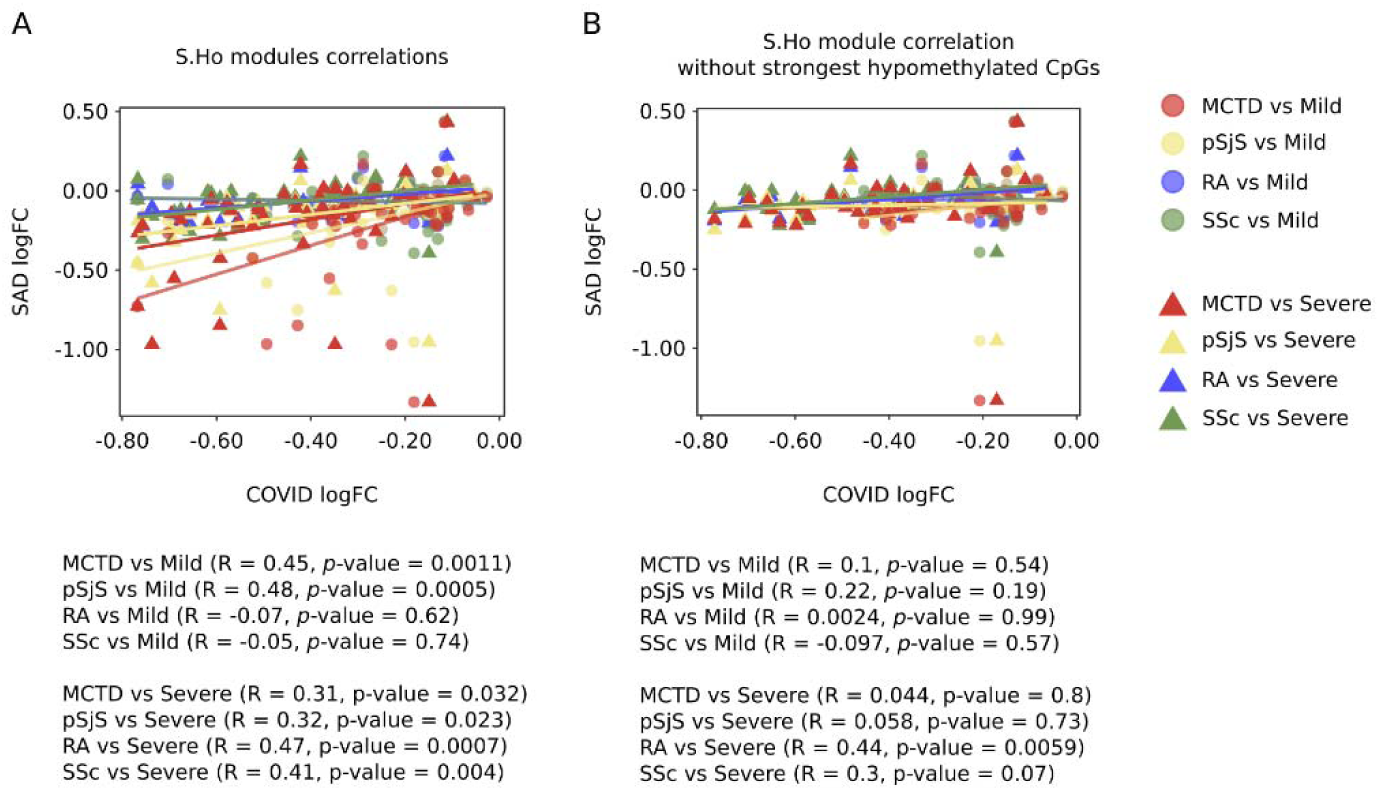
Progressive hypomethylation during COVID-19 severity CpG module (S.Ho) is composed of two different functional signatures. (A) logFC correlation plots for severe and mild COVID-19 cases against two interferon related diseases (MCTD and pSjS) and two non-interferon related diseases (RA and SSc) are shown. Correlation coefficients and *p*-values are shown by pairs. (B) logFC correlation plots without strongest hypomethylated CpGs are shown.

**Supplementary Figure 6:**
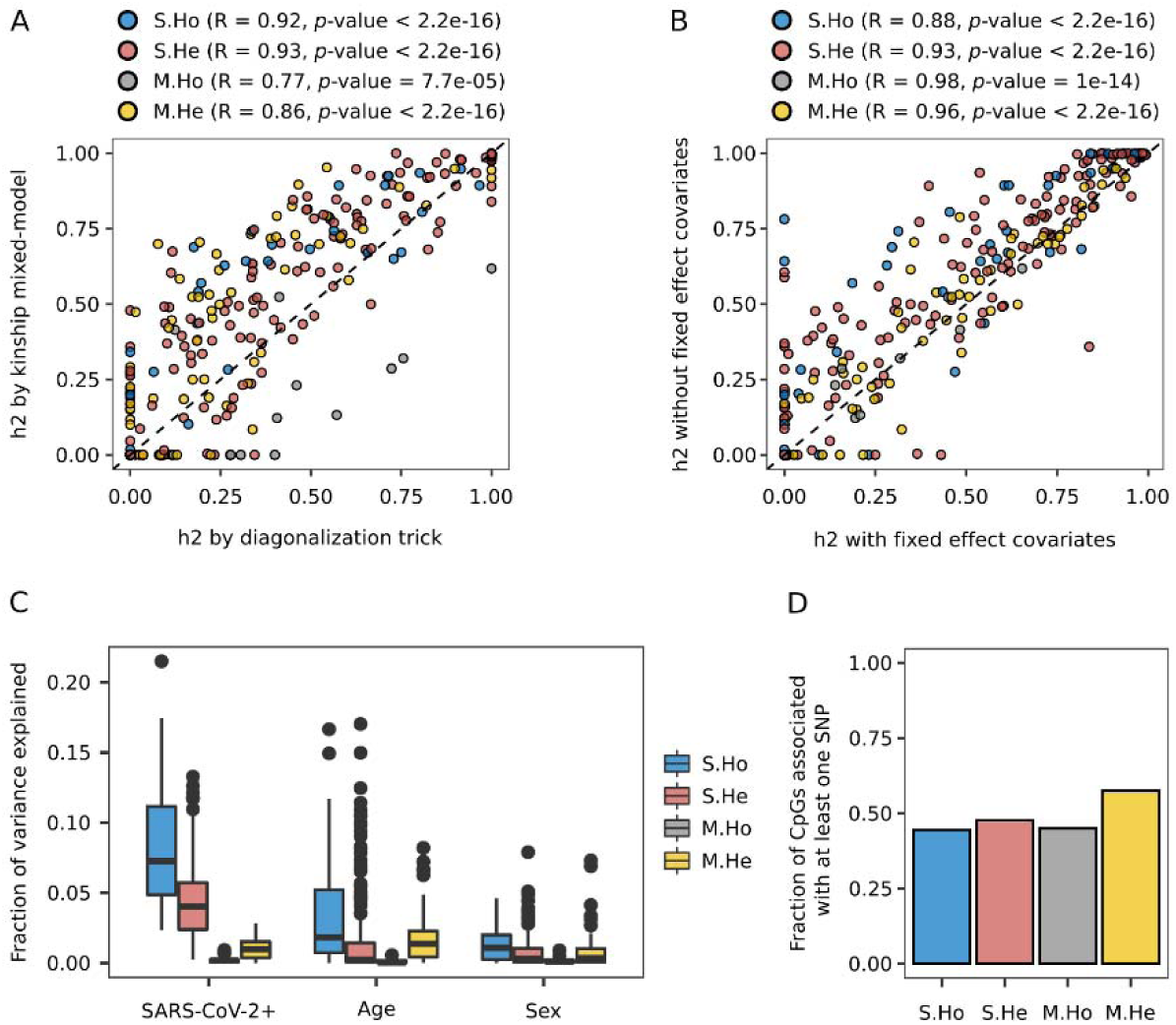
Genetic and non-genetic DNA methylation explained variance analyses. (A) Genetic heritability correlation between two independent methods is shown by CpG module (Linear mixed-model variance decomposition and Linear mixed-model fitting with the diagonalization trick were used). Correlation coefficients and *p*-values are depicted by module. (B) Genetic heritability correlation between linear mixed-model variance decomposition with and without fixed effect covariates (SARS-CoV-2 infection, gender and age) is plotted. Correlation coefficients and *p*-values are depicted by module. (C) Fraction of DNA methylation variance explained by fixed effect covariates is shown for each CpG module. (D) Fraction of CpGs by module associated with at least one SNP is shown.

